# Creating the Pick’s disease International Consortium: Association study of *MAPT* H2 haplotype with risk of Pick’s disease

**DOI:** 10.1101/2023.04.17.23288471

**Authors:** Rebecca R Valentino, William J Scotton, Shanu F Roemer, Tammaryn Lashley, Michael G Heckman, Maryam Shoai, Alejandro Martinez-Carrasco, Nicole Tamvaka, Ronald L Walton, Matthew C Baker, Hannah L Macpherson, Raquel Real, Alexandra I Soto-Beasley, Kin Mok, Tamas Revesz, Thomas T Warner, Zane Jaunmuktane, Bradley F Boeve, Elizabeth A Christopher, Michael DeTure, Ranjan Duara, Neill R Graff-Radford, Keith A Josephs, David S Knopman, Shunsuke Koga, Melissa E Murray, Kelly E Lyons, Rajesh Pahwa, Joseph E Parisi, Ronald C Petersen, Jennifer Whitwell, Lea T Grinberg, Bruce Miller, Athena Schlereth, William W Seeley, Salvatore Spina, Murray Grossman, David J Irwin, Edward B Lee, EunRan Suh, John Q Trojanowski, Vivianna M Van Deerlin, David A Wolk, Theresa R Connors, Patrick M Dooley, Matthew P Frosch, Derek H Oakley, Iban Aldecoa, Mircea Balasa, Ellen Gelpi, Sergi Borrego-Écija, Rosa Maria de Eugenio Huélamo, Jordi Gascon-Bayarri, Raquel Sánchez-Valle, Pilar Sanz-Cartagena, Gerard Piñol-Ripoll, Laura Molina-Porcel, Eileen H Bigio, Margaret E Flanagan, Tamar Gefen, Emily J Rogalski, Sandra Weintraub, Javier Redding-Ochoa, Koping Chang, Juan C Troncoso, Stefan Prokop, Kathy L Newell, Bernardino Ghetti, Matthew Jones, Anna Richardson, Andrew C Robinson, Federico Roncaroli, Julie Snowden, Kieren Allinson, Oliver Green, James B Rowe, Poonam Singh, Thomas G Beach, Geidy E Serrano, Xena E Flowers, James E Goldman, Allison C Heaps, Sandra P Leskinen, Andrew F Teich, Sandra E Black, Julia L Keith, Mario Masellis, Istvan Bodi, Andrew King, Safa-Al Sarraj, Claire Troakes, Glenda M Halliday, John R Hodges, Jillian J Kril, John B Kwok, Olivier Piguet, Marla Gearing, Thomas Arzberger, Sigrun Roeber, Johannes Attems, Christopher M Morris, Alan J Thomas, Bret M. Evers, Charles L White, Naguib Mechawar, Anne A Sieben, Patrick P Cras, Bart B De Vil, Peter Paul P.P. De Deyn, Charles Duyckaerts, Isabelle Le Ber, Danielle Seihean, Sabrina Turbant-Leclere, Ian R MacKenzie, Catriona McLean, Matthew D Cykowski, John F Ervin, Shih-Hsiu J Wang, Caroline Graff, Inger Nennesmo, Rashed M Nagra, James Riehl, Gabor G Kovacs, Giorgio Giaccone, Benedetta Nacmias, Manuela Neumann, Lee-Cyn Ang, Elizabeth C Finger, Cornelis Blauwendraat, Mike A Nalls, Andrew B Singleton, Dan Vitale, Cristina Cunha, Agostinho Carvalho, Zbigniew K Wszolek, Huw R Morris, Rosa Rademakers, John A Hardy, Dennis W Dickson, Jonathan D Rohrer, Owen A Ross

## Abstract

**Background:** Pick’s disease (PiD) is a rare and predominantly sporadic form of frontotemporal dementia that is classified as a primary tauopathy. PiD is pathologically defined by argyrophilic inclusion Pick bodies and ballooned neurons in the frontal and temporal brain lobes. PiD is characterised by the presence of Pick bodies which are formed from aggregated, hyperphosphorylated, 3-repeat tau proteins, encoded by the *MAPT* gene. The *MAPT* H2 haplotype has consistently been associated with a decreased disease risk of the 4-repeat tauopathies of progressive supranuclear palsy and corticobasal degeneration, however its role in susceptibility to PiD is unclear. The primary aim of this study was to evaluate the association between *MAPT* H2 and risk of PiD.

**Methods:** We established the Pick’s disease International Consortium (PIC) and collected 338 (60.7% male) pathologically confirmed PiD brains from 39 sites worldwide. 1,312 neurologically healthy clinical controls were recruited from Mayo Clinic Jacksonville, FL (N=881) or Rochester, MN (N=431). For the primary analysis, subjects were directly genotyped for *MAPT* H1-H2 haplotype-defining variant rs8070723. In secondary analysis, we genotyped and constructed the six-variant *MAPT* H1 subhaplotypes (rs1467967, rs242557, rs3785883, rs2471738, rs8070723, and rs7521).

**Findings:** Our primary analysis found that the *MAPT* H2 haplotype was associated with increased risk of PiD (OR: 1.35, 95% CI: 1.12-1.64 P=0.002). In secondary analysis involving H1 subhaplotypes, a protective association with PiD was observed for the H1f haplotype (0.0% vs. 1.2%, P=0.049), with a similar trend noted for H1b (OR: 0.76, 95% CI: 0.58-1.00, P=0.051). The 4-repeat tauopathy risk haplotype *MAPT* H1c was not associated with PiD susceptibility (OR: 0.93, 95% CI: 0.70-1.25, P=0.65).

**Interpretation:** The PIC represents the first opportunity to perform relatively large-scale studies to enhance our understanding of the pathobiology of PiD. This study demonstrates that in contrast to its protective role in 4R tauopathies, the *MAPT* H2 haplotype is associated with an increased risk of PiD. This finding is critical in directing isoform-related therapeutics for tauopathies.

**Funding:** See funding section

## Introduction

Pick’s disease (PiD) is a rare and predominantly sporadic subtype of frontotemporal lobar degeneration (FTLD) which represents approximately 5% of all dementias worldwide. Although there are no clinical diagnostic criteria for PiD, it typically develops in individuals approximately 55 years of age and presents with behavioral change, impaired cognition and occasionally motor difficulties (1-7). PiD is a relatively rapidly progressive disease and patients die approximately 10 years after disease onset (1-6). Symptomatic treatments are available, but currently no treatments exist that can delay disease onset or progression. A definite diagnosis of PiD requires autopsy confirmation.

Neuropathologically, PiD is classified by severe frontotemporal, knife-edge like cortical atrophy macroscopically, and microscopically the presence of ballooned neurons and argyrophilic, tau-immunoreactive inclusion “Pick bodies” in frontal and temporal regions (1). Characteristic Pick bodies consist of hyperphosphorylated 3-repeat (3R) tau aggregate proteins which are encoded by the *MAPT* gene on chromosome 17 (7, 8), and therefore PiD is classified as a 3R tauopathy. *MAPT* codes for six major tau isoforms in the adult human brain, and this is determined by alternative splicing of exons 2, 3, and 10 influencing the number of repeat domains in the N-terminus and C-terminus (9). More specifically, alternative splicing leading to exon 10 exclusion results in 3-repeat units in the microtubule binding C-terminal domain, generating 3R tau proteins (10).

Rare mutations in *MAPT* have been identified in a handful of PiD cases or individuals with PiD-like pathology (11-14); however, these data are inconsistent as larger, independent cohorts of PiD cases do not report *MAPT* mutations (15).The *MAPT* gene also has two well characterized common haplotypes, H1 and H2, which developed from a 900kb ancestral genetic inversion event (16). Not only has *MAPT* H1 consistently been associated with an increased risk of 4-repeat (4R) primary tauopathies such as progressive supranuclear palsy (PSP) and corticobasal degeneration (CBD), but the H1 haplotype is also the strongest genetic risk factor for both diseases (17, 18). To date, this observation has not been replicated in 3R tauopathy of PiD which may be due to the limited available sample size (19, 20).

Due to its rare prevalence and the inability to diagnose it clinically in life, PiD is an understudied neurodegenerative disease, and its genetic etiology is unknown. As previously mentioned, the few studies of *MAPT* haplotype in PiD that have been conducted were small and underpowered. Moreover, limited access to 3R tauopathy samples has stalled research advancement in understanding how *MAPT* haplotypes and isoforms influence disease risk/pathology and has prevented progress in developing isoform-specific therapies. To address this, we established the Pick’s disease International Consortium (PIC) and are collecting data from pathologically confirmed PiD cases from sites worldwide. Whilst also developing an in-depth consortium database of clinical, pathological, and demographic information, the primary aim of the PIC was to evaluate the association of the *MAPT* H1/H2 haplotype with disease risk, age of onset (AAO), and disease duration (DD) in PiD.

## Methods

### Pick’s disease International Consortium (PIC)

Due to the rare and understudied landscape of PiD, researchers at Mayo Clinic Brain Bank in Jacksonville, FL, USA (MC) and the UK Dementia Research Institute at University College London Queen Square Institute of Neurology (UCL) led efforts to establish the world’s first international consortium for Pick’s disease (PIC). MC led the effort for identifying and sourcing PiD cases from North American regions and UCL was responsible for collecting PiD cases from European and Australasian territories. Inclusion criteria were a neuropathologic diagnosis of PiD with Pick bodies and available frozen brain tissue. Exclusion criteria were frontotemporal dementia due to etiology other than a 3R predominant tauopathy or lack of frozen specimens. IRB approval was obtained for the studies at both collection hubs (MC and UCL) and each individual brain bank had institutional IRB approval for collection and sharing of specimens.

### Study Subjects

In the current study, 338 neuropathologically confirmed PiD cases were recruited from 39 sites worldwide (**Figure 1**), at the two major collection hubs in North America (MC) and Europe (UCL). Frozen brain tissue from cerebellum or prefrontal cortex were obtained from each case. All subjects were self-reported unrelated and Caucasian, non-Hispanic (genetically confirmed by array data). Baseline demographic information was collected for all subjects (AAO and age at death (AAD) for PiD patients, age at blood collection for controls, and sex). DD was calculated from the difference between AAD and AAO for a subset of 309 PiD cases. Subject characteristics are summarized in **Table 1**. In addition to basic demographic information, the PIC also collected information related to family histories, clinical outcomes (e.g. behavioral and language impairments, presence/absence of parkinsonism, upper and lower motor neurone deficits, Mini-Mental State Examination and Clinical Dementia Rating), and pathological information (e.g. Thal phase, Braak stage, and brain weight,) for each individual case, as well as noting whether other tissues and brain imaging data were available. Cases were removed if a rare *MAPT* mutation was identified. Peripheral blood-derived DNA was provided from 1,312 controls from Mayo Clinic in Jacksonville, FL (N=881) or Rochester, MN (N=431). Control subjects were deemed neurologically healthy by neurologists at Mayo Clinic.

**Figure 1.**
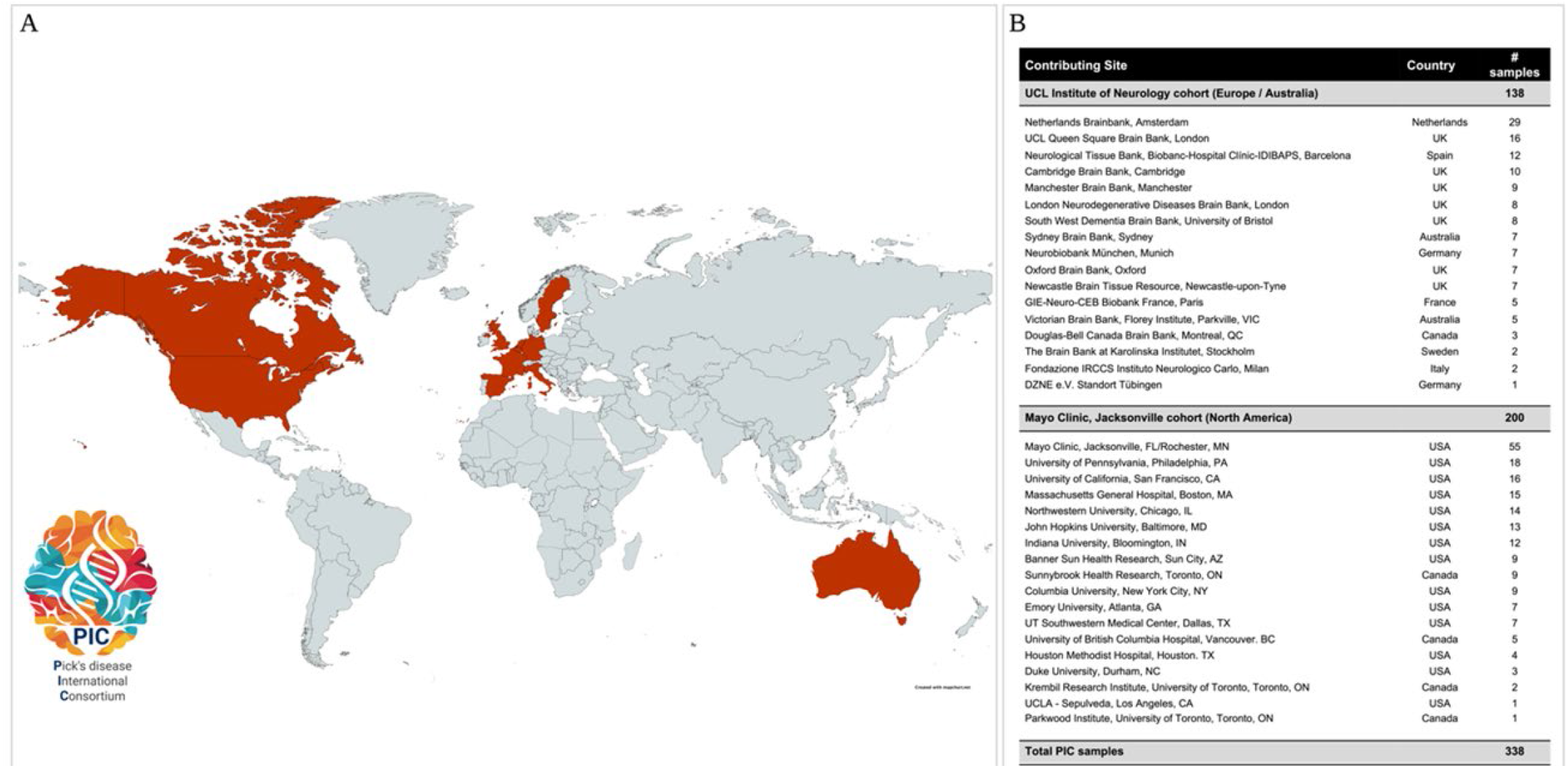
Global map and table reporting countries and recruitment sites that have contributed Pick’s disease tissues to the Pick’s disease International Consortium (PIC) to date. Dark red= countries that have collected and donated Pick’s disease tissues.

**Table 1:** Summary of subject characteristics. The sample median (minimum, maximum) is given for age. Age represents age at death in Pick’s disease cases and age at blood draw in controls. Age at disease onset and disease duration information was unavailable for N=29 Pick’s disease cases.

| Variable | Pick's disease series (N=338) | Controls (N=1,312) |
| --- | --- | --- |
| Age (years) | 69 (40, 100) | 69 (45, 92) |
| Age of disease onset (years) | 58 (33, 80) | N/A |
| Disease duration (years) | 10 (2, 25) | N/A |
| Sex |  |  |
| Male | 205 (60.7%) | 611 (46.6%) |
| Female | 133 (39.3%) | 701 (53.4%) |

### Established methods for the neuropathological diagnosis of Pick’s disease

Currently, diagnostic consensus criteria for the neuropathologic diagnosis of PiD do not exist. In many diagnostic centers a neuropathological diagnosis of PiD relies on the presence of argyrophilic, spherical neuronal inclusions using traditional silver staining methods, such as Bielschowsky’s or Gallyas-Braak silver staining methods. Both silver staining methods stain Alzheimer’s disease (AD) neurofibrillary tangles, yet spherical inclusions in PiD are positive with Bielschowsky and negative on the Gallyas-Braak silver staining method (21). This differentiation in silver staining methods is helpful especially for centers that do rely on immunohistochemistry against phosphorylated tau (p-tau) and do not have isotype specific tau antibodies incorporated in the diagnostic work-up as AD and PiD neuropathologic changes may co-exist in the same patient. Immunohistochemistry against epitope-specific tau antibodies further helps to distinguish between AD and PiD features. Since both spherical inclusions and neurofibrillary tangles stain positive with antibodies against phosphorylated tau (p-tau), epitope-specific antibodies highlight selective 3R tau spherical inclusions in PiD, which is further validated by antibodies to 4R tau where these spherical inclusions stain negative. This distinction is particularly obvious in the granule cell neurons of the hippocampal dentate fascia, which may be used solely to diagnose PiD.

### PIC diagnostic algorithm for pathology confirmed Pick’s disease

Since a harmonized neuropathologic diagnostic scheme does not exist it became pivotal to the PIC aims to establish a defined set of operational diagnostic criteria within PIC that would ensure that submitted PiD cases reflect a 3R-predominant tauopathy. All cases submitted to the PIC had an archival neuropathologic diagnosis of PiD (i.e. the presence of argyrophilic or p-tau positive spherical inclusions) and underwent neuropathological assessments at their respective brain banks. Due to the multi-site nature of the PIC, each participating center were requested to submit and report respective 3R and 4R tau staining results for each individual PiD case to the PIC. To fulfill PIC criteria all cases had to confirm the presence of Pick bodies and must have had 3R tau positive and 4R tau negative inclusions. The additional presence of ballooned neurons and negative Gallyas staining of inclusions was preferred to confirm diagnosis. If 3R/4R tau immunohistochemistry had not been performed at their respective brain banks, centers submitted routinely cut sections (up to seven microns) of unstained, formalin fixed paraffin embedded tissue from hippocampal, frontal or temporal lobe regions for 3R and 4R tau immunohistochemistry assessments (**Figure 2**). Cases submitted to Mayo Clinic Brain Bank for Neurodegenerative Diseases were evaluated by two PIC neuropathologists (DWD, SFR) and cases submitted to UCL were examined by two PIC investigators (WS, TL) which included a PIC neuropathologist (TL), all using the PIC diagnostic algorithm. All sections were stained using standard immunohistochemical methods (22) (**Figure 3**).

**Figure 2.**
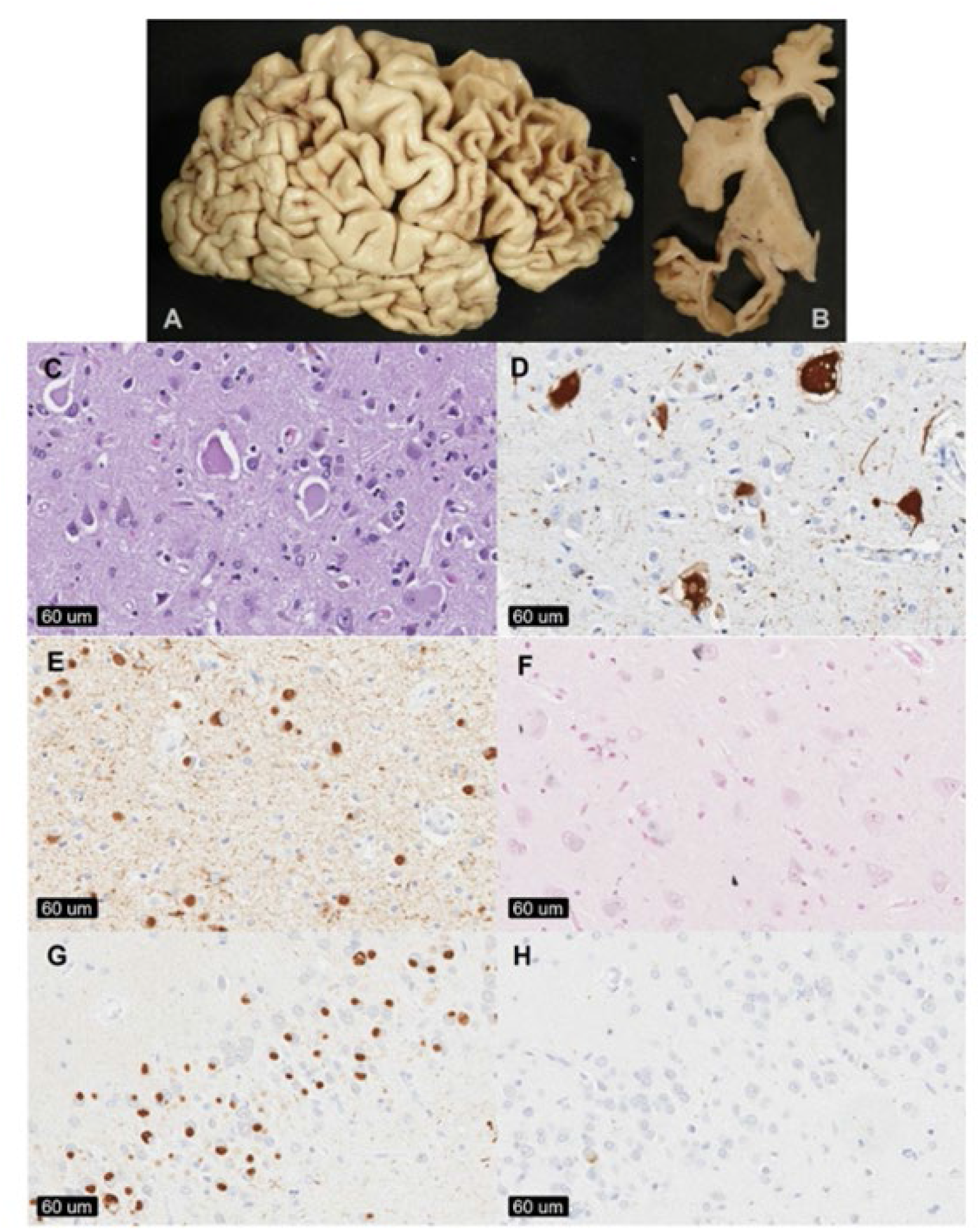
Pothologicnt assessments of Pick’s diseose brains ot Mayo Clinic Brain Bank for Neurodegenerative Diseases in jaeksonviUe, FL, LS4. (A) The superior and dorsoioteroi surfaces of the frontal cortex ano tempore! tobe often show severe circumscribed ‘knife-edge’ edge atrophy [Bj Coronal sections of the brain show markedly dilated ventricles, cortical atrophy, and hippocampal affection. [C] Enlarged, amorphous ballooned neurons. [D] in regions with severe astrogliosis and neuronal loss, iioinjrtg o^ojrijr aS-iiysraiirrt may highlight ballooned neurons. [E] Phosphorylated tau antibodies highlight spherical cytoplasmic neuronal inclusions and may also show marked neuropil staining, especially in cases with concomitant Alzheimer’s type pathology. [F] Galiyas situer stains moy stoin isolated glial lesions or neurofibrillary tangles; however, Pick bodies do not show any significant degree of silver staining. [G] 3 ft tau staining of tie tfenttrte fascia of the hippocampus show strong immunoreoctivity of spherical inclusions. [H] 4R tau staining of the dentate fascia show negative spherical inclusion, although isolated neurofibrillary tangles may siorii positive. Images ore from Pick coses submitted to MCJ./

**Figure 3.**
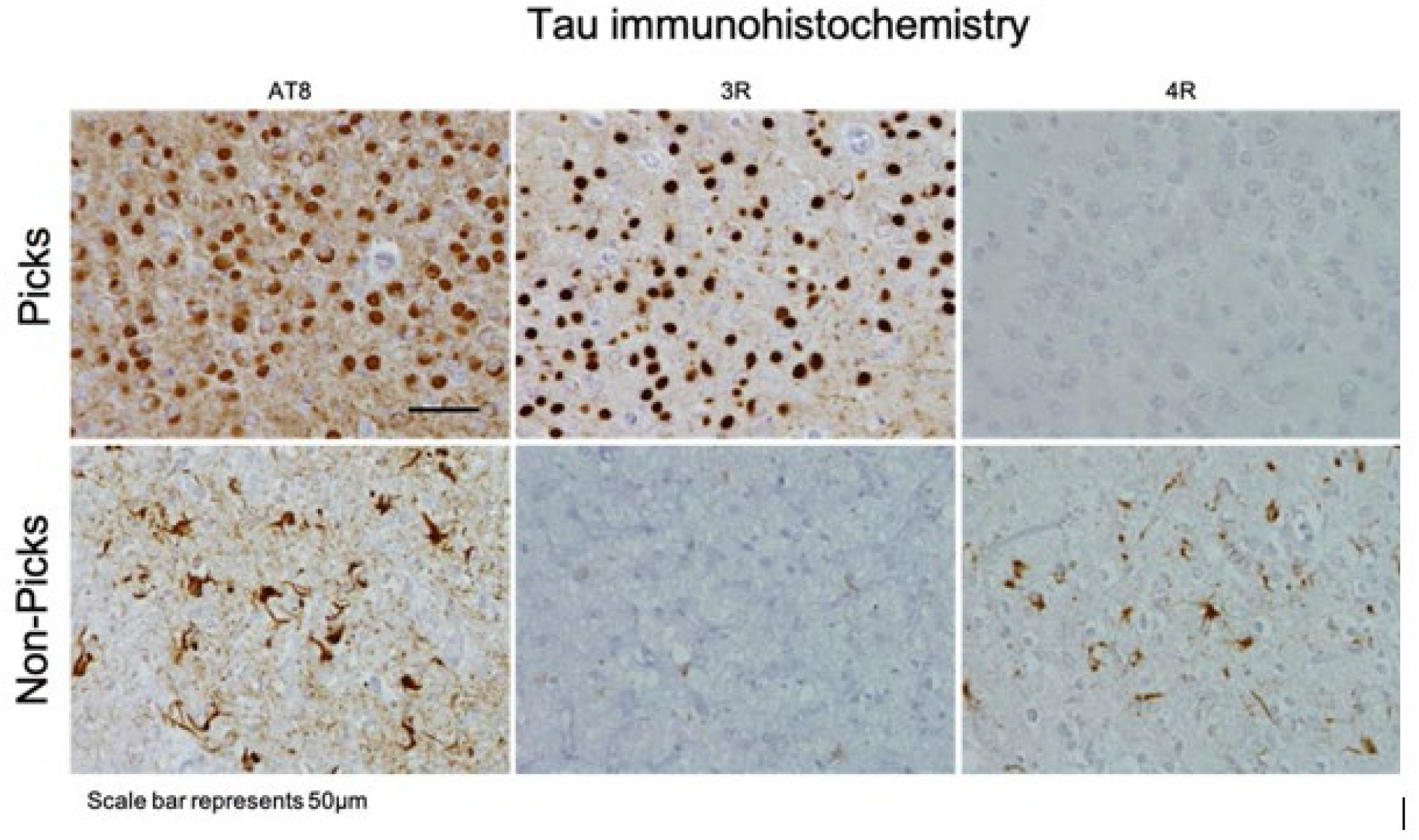
Pothologica! ossessments of Pick’s diseose brains ot Queen Square Brain Bank for Neurological Disorders (QSBB), UCL Queen Square Institute of Neurology, London, UK. The top row shows a Pick’s disease case that wos positive for AT8 and 3R-tau immunoreactive Pick bodies. The bottom row shows o non-Pick’s diseose case (that wos originally pathologically diagnosed with Pick’s disease) thot was positive for ATS and 4R-tou but negative for 3R tau immunoreactive Pick bodies, images are from Pick cases submitted to UCL.

### DNA Preparation

DNA was extracted from each subject at their respective collection site. At MC, genomic DNA was extracted from frozen brain tissue from PiD cases and from peripheral blood lymphocytes from control subjects using an automated or manual method. Automated DNA extractions were carried out using Autogen Tissue Kit reagents according to manufacturer protocols and were processed on the Autogen FlexSTAR+ (both Autogen, Holliston, MA, USA). At QSBB, total genomic DNA was extracted from frozen brain tissue using the Kleargene XL Nucleic Acid Purification kit (LGC, Hoddesdon, Herts, UK). DNA quality was assessed with a NanoDrop 8000 spectrophotometer (ThermoFisher Scientific, USA) and absorbance ratios for 260/280 and 260/230 were between 1.7-2.2 and 2.0-2.2, respectively.

### SNP Genotyping

The *MAPT* H2 haplotype-tagging variant rs8070723 was genotyped in all cases and controls. In addition, the five common *MAPT* variants (rs1467967, rs242557 [the H1C haplotype-tagging variant], rs3785883, rs2471738, and rs7521) which along with rs8070723 define H1-subhaplotypes were genotyped to assess *MAPT* subhaplotype structure (23, 24). North American cases and all controls were genotyped using TaqMan SNP genotyping assays on an ABI 7900HT Fast Real-Time PCR system (Applied Bio-systems, Foster City, CA, USA), as previously described (25). *MAPT* variants were genotyped according to manufacturer instructions (primer sequences available upon request). Genotypes were called using TaqMan Genotyper Software v1.3 (Applied Bio-systems, Foster City, CA, USA). European and Australasian cases were genotyped using KASP™ SNP genotyping assays on the Hydrocyler2 system (LGC Genomics, Hoddesdon, Herts, UK) according to manufacturer instructions, and were read on a PHERAStar FSX plate reader (BMG Labtech, Cary, NC, USA). Genotypes were called using Kraken KlusterKaller™ software (LGC Genomics, Hoddesdon, Herts, UK). Genotype call rates for all subjects were 100% for each variant. There was no evidence of a departure from Hardy-Weinberg equilibrium in controls for any of the six variants (all P >0.01 after Bonferroni correction). All cases were assessed for population stratification using available whole SNP genotyping data. After standard genotyping data quality control steps, we performed a principal components analysis (PCA), merged all cases with the European (CEU) HapMap reference dataset, and identified any cases of non-white European ancestry which were excluded from further analysis. Allele and genotype frequencies for each variant are detailed in **Supplementary Table 1**.

### Statistical Analysis

Single-variant associations with risk of PiD were evaluated using logistic regression models that were adjusted for age and sex. Odds ratios (Ors) and 95% confidence intervals (Cis) were estimated and correspond to each additional minor allele. Single-variant associations with AAO and DD in PiD patients were examined using linear regression models that were adjusted for sex and series (AAO analysis) or sex, AAO, and series (DD analysis). Regression coefficients (referred to as β) and 95% Cis were estimated and are interpreted as the increase in the mean AAO or DD corresponding to each additional copy of the minor allele. For all single-variant associations, analysis involving rs8070723 (the H2-tagging variant) was considered as the primary analysis, with results for the five remaining variants considered as secondary and presented for completeness.

Associations between six-variant *MAPT* haplotypes and risk of PiD were assessed using score tests for association under a logistic regression framework (26), where tests were adjusted for age and sex. Ors and 95% Cis were estimated and correspond to each additional copy of the given haplotype.

In analysis of PiD patients, associations of six-variant *MAPT* haplotypes with AAO and DD were assessed using score tests for association under a linear regression framework (26), where tests were adjusted for sex and series (AAO analysis) or sex, AAO, and series (DD analysis). B-coefficients and 95% Cis were estimated and are interpreted as the increase in the mean AAO or DD corresponding to each additional copy of the given haplotype. Haplotypes occurring in <1% of subjects in a specific analysis were excluded from that analysis.

We adjusted for multiple testing separately for each outcome measure that was examined (presence of PiD, AAO, or DD). P-values <0.05 were considered as statistically significant in the primary analysis involving the *MAPT* rs8070723 variant. In secondary analysis assessing associations between *MAPT* haplotypes and outcomes, p-values < 0.0028 (18 tests) were considered as statistically significant after Bonferroni correction in the disease-association analysis, and p-values < 0.0031 (16 tests) were considered as statistically significant in the AAO and DD analyses. P-values ≤ 0.05 were considered as significant in all remaining analysis. All statistical tests were two-sided. Statistical analyses were performed using R Statistical Software (version 4.1.2; R Foundation for Statistical Computing, Vienna, Austria).

## Results

A total of 338 pathologic-defined PiD cases were identified across 39 independent recruitment sites to establish the first PiD consortium (PIC). There was a significant association between the *MAPT* rs8070723 H2 allele and an increased risk of PiD in the overall series (OR: 1.35, 95% CI: 1.12-1.64, P=0.0021), with minor allele frequencies of 29.0% in the 338 PiD patients and 23.0% in the 1,312 controls. *MAPT* rs8070723 was not associated with AAO (β: -0.54, 95% CI: -1.94 to 0.87, P=0.45) or DD (β: 0.25, 95% CI: -0.46 to 0.96, P=0.50). Single-variant associations with PiD, AAO and DD are shown for all six *MAPT* variants used to define *MAPT* haplotypes in **Supplementary Tables 2 and 3**. Of note, there was not a notable association between rs242557 and risk of PiD (OR: 0.94, 95% CI: 0.79-1.12, P=0.51, **Supplementary Table 2**).

In secondary analysis, an evaluation of associations between six-variant *MAPT* haplotypes and risk of PiD is displayed in **Table 2**. As with the single-variant analysis, the H2 haplotype was associated with an increased risk of PiD (OR: 1.34, 95% CI:1.11-1.63, P=0.0028); the slight difference between the two numerical estimates is due to the two different analysis approaches. Additionally, a nominally significant (P<0.05) protective association was noted for the rare H1f haplotype (0.0% in PiD, 1.2% in controls, P=0.049), with a slightly weaker finding noted for H1b (OR: 0.76, 95% CI: 0.58-1.00, P=0.051). There were no other notable associations between *MAPT* haplotypes and risk of PiD (all P≥0.15, **Table 2**).

**Table 2:** Associations between MAPT haplotypes and risk of Pick’s disease. ORs, 95% CIs, and p-values result from score tests of association that were adjusted for age and sex. 1Indicates a haplotype that was not observed in Pick’s disease patients, making estimation of an OR impossible. P-values <0.0028 are considered as statistically significant after applying a Bonferroni correction for multiple testing. OR=odds ratio; CI=confidence interval.

| Haplotype | MAPT variant |  |  |  |  |  | Haplotype frequency (%) |  | Association with Pick's disease |  |
| --- | --- | --- | --- | --- | --- | --- | --- | --- | --- | --- |
|  | rs1467967 | rs242557 | rs3785883 | rs2471738 | rs8070723 | rs7521 | Pick's disease patients (N=338) | Controls (N=1312) | OR (95% CI) | P-value |
| H1b | G | G | G | C | A | A | 13.1 | 16.0 | 0.76 (0.58, 1.00) | 0.051 |
| H1c | A | A | G | T | A | G | 10.2 | 11.3 | 0.93 (0.70, 1.25) | 0.65 |
| H1d | A | A | G | C | A | A | 7.4 | 7.1 | 0.99 (0.68, 1.42) | 0.94 |
| H1e | A | G | G | C | A | A | 9.8 | 9.0 | 1.03 (0.74, 1.42) | 0.87 |
| H1f | G | G | A | C | A | A | 0.0 | 1.2 | N/A<sup>1</sup> | 0.049 |
| H1g | G | A | A | C | A | A | 0.7 | 1.1 | 0.43 (0.11, 1.65) | 0.22 |
| H1h | A | G | A | C | A | A | 4.0 | 4.1 | 0.95 (0.57, 1.57) | 0.85 |
| H1i | G | A | G | C | A | A | 3.9 | 4.4 | 0.98 (0.60, 1.61) | 0.95 |
| H1l | A | G | A | C | A | G | 3.6 | 3.0 | 1.11 (0.67, 1.84) | 0.69 |
| H1m | G | A | G | C | A | G | 2.9 | 2.9 | 1.00 (0.56, 1.78) | 0.99 |
| H1o | A | A | A | C | A | A | 1.1 | 2.3 | 0.53 (0.23, 1.26) | 0.15 |
| H1p | G | G | G | T | A | G | 1.1 | 1.5 | 0.82 (0.33, 2.04) | 0.66 |
| H1r | A | G | G | T | A | G | 0.7 | 1.1 | 0.63 (0.20, 2.01) | 0.44 |
| H1u | A | A | G | C | A | G | 2.4 | 2.4 | 1.11 (0.58, 2.11) | 0.75 |
| H1v | G | G | A | T | A | G | 2.2 | 1.2 | 1.50 (0.70, 3.21) | 0.30 |
| H1x | G | A | A | T | A | G | 1.3 | 1.3 | 1.06 (0.44, 2.56) | 0.91 |
| H1y | A | A | A | T | A | G | 1.4 | 1.6 | 0.85 (0.34, 2.07) | 0.71 |
| H2 | A | G | G | C | G | G | 28.5 | 22.7 | 1.34 (1.11, 1.63) | 0.0028 |

Associations of *MAPT* haplotypes with AAO and DD in PiD patients are shown in **Table 3**. None of the six-variant *MAPT* haplotypes were significantly associated with AAO or DD after correcting for multiple testing (P<0.0031 considered significant). However, nominally significant associations were observed with AAO for H1b (β: 2.66, 95% CI: 0.63 to 4.70, P=0.011), H1i (β: -3.66, 95% CI: -6.83 to - 0.48, P=0.025) and H1u (β: -5.25, 95% CI: -10.42 to -0.07, P=0.048), and with a shorter DD for H1x (β: -3.73, 95% CI: -6.98 to -0.48, P=0.025).

**Table 3:** Associations of MAPT haplotype with age of disease onset and disease duration in Pick’s disease cases. β values, 95% CIs, and p-values result from score tests of association that were adjusted for sex and series (age of disease onset analysis) or sex, age of disease onset, and series (disease duration analysis). β values are interpreted as the change in the mean value of the given outcome (age of disease onset or disease duration) corresponding to each additional copy of the given haplotype. P-values <0.0031 are considered as statistically significant after applying a Bonferroni correction for multiple testing. β=regression coefficient; CI=confidence interval.

| Haplotype | Haplotype frequency (%), N=309 | Association with age of disease onset |  | Association with disease duration |  |
| --- | --- | --- | --- | --- | --- |
| | | $\beta$ (95% CI) | P-value | $\beta$ (95% CI) | P-value |
| H1b | 13.3% | 2.66 (0.63, 4.70) | 0.011 | -0.03 (-1.07, 1.02) | 0.96 |
| H1c | 10.0% | 1.63 (-0.61, 3.86) | 0.15 | 0.08 (-1.05, 1.22) | 0.88 |
| H1d | 7.2% | 0.79 (-1.79, 3.38) | 0.55 | -0.91 (-2.21, 0.39) | 0.17 |
| H1e | 9.3% | 0.52 (-1.94, 2.98) | 0.68 | 0.52 (-0.72, 1.76) | 0.41 |
| H1h | 4.0% | 2.03 (-1.57, 5.64) | 0.27 | -0.45 (-2.27, 1.37) | 0.63 |
| H1i | 4.1% | -3.66 (-6.83, -0.48) | 0.025 | -0.90 (-2.53, 0.72) | 0.28 |
| H1l | 3.5% | -1.75 (-5.42, 1.92) | 0.35 | 0.43 (-1.42, 2.28) | 0.65 |
| H1m | 3.1% | -1.25 (-5.33, 2.84) | 0.55 | 0.94 (-1.11, 3.00) | 0.37 |
| H1o | 1.2% | 0.05 (-6.91, 7.00) | 0.99 | 0.03 (-3.47, 3.52) | 0.99 |
| H1p | 1.0% | -5.65 (-12.60, 1.30) | 0.11 | 0.17 (-3.36, 3.69) | 0.93 |
| H1u | 2.2% | -5.25 (-10.42, -0.07) | 0.048 | -2.40 (-5.03, 0.22) | 0.074 |
| H1v | 2.1% | -1.74 (-6.61, 3.13) | 0.48 | 1.91 (-0.54, 4.35) | 0.13 |
| H1x | 1.4% | -5.39 (-11.84, 1.07) | 0.10 | -3.73 (-6.98, -0.48) | 0.025 |
| H1y | 1.5% | -0.70 (-6.93, 5.54) | 0.83 | 1.82 (-1.31, 4.95) | 0.26 |
| H1z | 1.6% | -1.81 (-8.02, 4.40) | 0.57 | -0.08 (-3.20, 3.05) | 0.96 |
| H2 | 29.4% | -0.62 (-2.03, 0.79) | 0.39 | 0.22 (-0.49, 0.93) | 0.54 |

## Discussion

PiD is a rare, predominantly sporadic 3R tauopathy that presents primarily as a behavioral or language variant of frontotemporal dementia (1-6). Little is known regarding the etiology or underlying pathobiology of the disease. To date, no genetic variation has been shown to associate with disease risk, although three cases with PiD or PiD-like pathology have been suggested to be caused by rare *MAPT* mutations (11-14). In the present study we have shown that the common *MAPT* H2 haplotype, strongly protective in 4R-tauopathy, is associated with an increased risk of PiD (3R tauopathy). This was only possible by establishing and creating a global consortium (PIC) to increase the number of available pathologically-defined PiD cases. Previous early genetic studies were underpowered with only 34 cases and 33 cases respectively (19, 20); a ten-fold increase in sample size was needed to establish *MAPT* H2 as a risk factor for in PiD.

Previous research in frontotemporal dementia linked to chromosome 17 with tau pathology (FTDP17t) has clearly demonstrated that mutations in the 5′ splice site of *MAPT* exon 10 can increase the incorporation of the exon into the mRNA and increase 4R isoform production, emphasizing how important exon 10 alternative splicing regulation is as dysregulation influences tangle formation and neurodegeneration outcome (16, 27). Given the association of *MAPT* H2 with a 3R-tauopathy, and its protection in 4R-tauopathy, it is possible that the *MAPT* H1 and H2 haplotypes increase the expression of 4R and 3R tau respectively. Previous studies have attempted to investigate the haplotype influence on *MAPT*/tau expression although results have been inconclusive, given the presence of six different isoforms in human brain defining specific isoform expression remains complex (28-30). The genetic predisposition herein described would appear to support the hypothesis that the pathologic effects of the H1-H2 haplotypes is via isoform specific expression differences. This may have implications in the determination of therapeutic strategies that have focused on either overall lowering of tau expression or specifically targeting the lowering of 4R-tau or increasing 3R-tau isoforms. The overall balance of the 3R and 4R forms of tau would appear to be important for the primary tauopathies but does not in itself explain the mixed pathology observed in AD, although it is tempting to suggest an overall increased expression of total tau may be underlying the mixed pathology.

In addition to providing evidence that the *MAPT*-H2 haplotype is associated with an increased risk of PiD, we observed nominally significant associations that were observed with risk of PiD, AAO, and DD, however these will require validation. This study has strengths in the assembled large PiD series of patients and the direct genotyping of the H1-H2 haplotype, but there still remains several limitations which are important to note. The possibility of a type II error (i.e. false-negative finding) is important to consider, and we cannot conclude that there is no true association between a given haplotype and risk of PiD simply due to a non-significant p-value in this study. Additionally, we were unable to regress out genetic principal components, and so it is possible that population stratification could have had an influence on our results. However, we used the case genetic principal components to exclude any cases of non-European ancestry and our control MAPT H1-H2 frequencies were in keeping with published data and the general population frequency, in fact the highest population control frequency in gnomAD is 23.8% very similar to our 23% (31). Ongoing studies looking at genome-wide disease associations in PiD with available genome-wide SNP data for controls support the current findings (data not shown).

In summary, PiD is a rare and understudied disease with a devastating impact on both patients and their families. Through collaboration and building of the PIC, we have for the first time a rare opportunity to engage in studies that may tease out the underlying pathobiology in PiD. As a primary tauopathy, there is the possibility that the identification of genetic variables, such as *MAPT* H2, involved in PiD pathology will inform on other more common tau-related disorders, PSP, CBD, and potentially AD. Larger scale unbiased studies to explore genome-wide or structural genetic variation in PiD are now warranted. Furthermore, resolving the genetic determinants of PiD may help in establishing diagnostic criteria and elucidating the dysfunctional pathways may direct future therapeutic intervention strategies.

## Data Availability

The PIC have built a database that contains detailed demographic, clinical, and pathological information for deidentified participants with Picks disease. Basic demographic information (e.g. age at onset, age at death, disease duration, sex, and ethnicity), family history, clinical history (e.g. behavioral and language impairments, presence of parkinsonism, upper and lower motor deficits, MMSE, and CDR), and pathological observations (e.g. immunohistochemical staining records, Thal phase, Braak stage, TDP-43 type, post-mortem intervals, brain weight, and vascular pathology), other available tissues, genetic data and clinical imaging data are available for each subject upon request. All requests must be submitted to Owen A. Ross or Jonathan Rohrer.

## Data sharing

The PIC have built a database that contains detailed demographic, clinical, and pathological information for deidentified participants with Pick’s disease. Basic demographic information (e.g. age at onset, age at death, disease duration, sex, and ethnicity), family history, clinical history (e.g. behavioral and language impairments, presence of parkinsonism, upper and lower motor deficits, MMSE, and CDR), and pathological observations (e.g. immunohistochemical staining records, Thal phase, Braak stage, TDP-43 type, post-mortem intervals, brain weight, and vascular pathology), other available tissues, genetic data and clinical imaging data are available for each subject upon request. All requests must be submitted to Owen A. Ross or Jonathan Rohrer.

## Acknowledgements

This paper is dedicated to the memory of John Q. Trojanowski, M.D. Ph.D., who was an inspirational researcher and neuropathologist at the University of Pennsylvania and pioneered discoveries in tauopathies that resulted in improvements of diagnosis and available treatments. John was a leader in neuroscience and his presence and insights will be thoroughly missed by scientists worldwide. We would also like to acknowledge our dear colleagues Charles Duyckaerts and Murray Grossman, eminent neuropathologists at Sorbonne University, Paris and University of Pennsylvania, USA. Charles Duyckaerts sadly passed away during our manuscript writing after a long battle with cancer. We sincerely thank all those who contributed towards our research, particularly the patients and families who donated brain and blood tissues. Without their generous donations the PIC would not exist, and this study would not have been possible. We also thank Audrey Strongosky for processing research subjects’ consents, drawing bloods, and handling collection procedures, as well as Linda G. Rousseau, Virginia R. Phillips, and Monica Castanedes‐Casey for their continuous commitment, technical support, and teamwork.

*Mayo Clinic:* SK receives funding from CurePSP and the Rainwater Charitable Foundation, the State of Florida Ed and Ethel Moore Alzheimer’s Disease Research Program (22A05), and Mayo Clinic Alzheimer’s Disease Research Center (ADRC).. MEM receives funding from the State of Florida (20A22), LEADS Neuropathology Core (U01AG057195), and the Chan Zuckerberg Initiative Collaborative Pairs Grant, which are paid directly to the institute. KAJ is supported by National Institutes of Health (NIH) grants (R01 DC014942, R01, R01-AG37491, R01-NS89757, RF1-NS112153 and RF1-NS120992). BFB is supported by National Institutes of Health (NIH) grants (P30 AG62677, U19 AG063911, U01 NS100620, U19 AG071754); the Robert H. and Clarice Smith and Abigail Van Buren Alzheimer s Disease Research Program of the Mayo Foundation; the Lewy Body Dementia Association; the Mayo Clinic Dorothy and Harry T. Mangurian Jr. Lewy Body Dementia Program; the Little Family Foundation; the Turner Family Foundation. ZKW is partially supported by the NIH/NIA and NIH/NINDS (1U19AG063911, FAIN: U19AG063911), Mayo Clinic Center for Regenerative Medicine, the gifts from the Donald G. and Jodi P. Heeringa Family, the Haworth Family Professorship in Neurodegenerative Diseases fund, and The Albertson Parkinson’s Research Foundation. He serves as PI or Co-PI on Biohaven Pharmaceuticals, Inc. (BHV4157-206), Neuraly, Inc. (NLY01-PD-1), and Vigil Neuroscience, Inc. (VGL101-01.002, VGL101-01.201, PET tracer development protocol, and Csf1r biomarker and repository project) grants. He serves as Co-PI of the Mayo Clinic APDA Center for Advanced Research and as an external advisory board member for the Vigil Neuroscience, Inc. OAR and DWD are both supported by National Institute of Neurological Disorders and Stroke (NINDS) Tau Center without Walls Program (U54-NS100693) and NIH (UG3-NS104095). DWD receives research support from the NIH (P30 AG062677; U54-NS100693; P01-AG003949), CurePSP, the Tau Consortium, and the Robert E. Jacoby Professorship. OAR is supported by NIH (P50-NS072187; R01-NS078086; U54-NS100693; U54-NS110435), Department of Defence (DOD) (W81XWH-17-1-0249) The Michael J. Fox Foundation, The Little Family Foundation, the Mangurian Foundation Lewy Body Dementia Program at Mayo Clinic, the Turner Family Foundation, Mayo Clinic Foundation, and the Center for Individualized Medicine. Mayo Clinic is also an LBD Center without Walls (U54-NS110435). Samples included in this study were clinical controls from Mayo Clinic Rochester and Mayo Clinic Jacksonville as part of the Alzheimer’s Disease Research Center (P30 AG062677), and the Mayo Clinic Study of Aging (U01 AG006786) or tissue donations to the Mayo Clinic Brain Bank in Jacksonville which is supported by CurePSP and Mayo Clinic funding.

*UCSF*: Human tissue samples were provided by the Neurodegenerative Disease Brain Bank at the University of California, San Francisco, which receives funding support from NIH grants P01AG019724 and P50AG023501, the Consortium for Frontotemporal Dementia Research, and the Tau Consortium. LTG and SS receive funding from NIH grants K24053435 and K08AG052648, respectively.

*UPenn*: ES, JQT, MG, VMVD, DJI, DAW, and EBL all receive funding through NIH - ES: P01-AG017586, P01-AG066597, P30-AG010124, P30-AG072979; JQT: P01-AG017586, P30-AG010124, P30-AG072979; MG: P01-AG017586, P01-AG066597, P30-AG010124, P30-AG072979; VMVD: P01-AG017586, P01-AG066597, P30-AG010124, P30-AG072979; DJI: R01-NS109260, P30-AG010124, P01-AG066597; DAW: P30-AG010124, P30-AG072979; and EBL: P01-AG017586, P01-AG066597, P30-AG010124, P30-AG072979.

*Northwestern*: ER, TG, SW, EHB, MEF) receive support from NIA under award numbers R01 AG062566, R01 AG077444, P30 AG13854, P30 AG072977; the National Institute of Deafness and Other Communication Disorders (NIDCD) under award number R01 DC008552; and the National Institute of Neurological Disorders and Stroke (NINDS) under award number R01 NS075075. MEF also receives support from NIA grant K08 AG065463.

*Banner*: We are grateful to the Banner Sun Health Research Institute Brain and Body Donation Program of Sun City, Arizona for the provision of human biological materials. The Brain and Body Donation Program has been supported by the NINDS (U24 NS072026 National Brain and Tissue Resource for Parkinson’s Disease and Related Disorders), the NIA (P30 AG19610 Arizona Alzheimer’s Disease Core Center), the Arizona Department of Health Services (contract 211002, Arizona Alzheimer’s Research Center), the Arizona Biomedical Research Commission (contracts 4001, 0011, 05-901 and 1001 to the Arizona Parkinson’s Disease Consortium), and the Michael J. Fox Foundation for Parkinson’s Research.

*Emory*: MG is supported by NIH grant P30 AG066511. MDC receives funding from NIH grant RF1 NS118584.

*Columbia*: We thank the Columbia University Alzheimer’s Disease Research Center (ADRC), funded by NIH grant P30AG066462, to S.A. Small (P.I.), and A. Teich from New York Brain Bank for providing biological samples and associated information. The ADRC is supported by the National Institutes of Health, through grant number P30AG066462. ACH receives NIH support through P30AG066462.

*Duke:* The Bryan Brain Bank and Biorepository of the Duke-UNC ADRC and SJW are supported by the NIA grant P30AG072958

*MGH:* Brain samples were provided by Neuropathology Core of the Massachusetts Alzheimer Disease Research Center, which receives funding support from NIH grant P30 AG062421, which also supported TRC, PMD, MPF and DHO. DHO was also received support from the Dr. and Mrs. E. P. Richardson, Jr, Fellowship in Neuropathology.

*Indiana:* BG is supported by the US National Institutes of Health (grant P30-AG010133).

*NIH:* This research was supported in part by the Intramural Research Program of the NIH, National Institute on Aging (NIA), National Institutes of Health, Department of Health and Human Services; project number ZO1 AG000535, as well as the National Institute of Neurological Disorders and Stroke. This work utilized the computational resources of the NIH HPC Biowulf cluster. (http://hpc.nih.gov).

*Western University*: We also acknowledge the DEC Brain & Biobank.

*Krembil*: GGK receives funding from The Rossy Foundation and Edmond J. Safra Philanthropic Foundation.

*McGill*: The Douglas-Bell Canada Brain Bank is funded by Healthy Brains for Healthy Lives (CFREF), the Réseau Québécois sur le suicide, le troubles de l’humeur et les troubles associés (FRQ-S), and by Brain Canada. NM is funded by a CIHR project grant.

*UCL*: WJS is a Wellcome Trust Clinical PhD Fellowship (220582/Z/20/Z). JDR is a Miriam Marks Brain Research UK Senior Fellowship and has received funding from an MRC Clinician Scientist Fellowship (MR/M008525/1) and the NIHR Rare Disease Translational Research Collaboration (BRC149/NS/MH). TL receives an Alzheimer’s Research UK senior fellowship. HRM is supported by research grants from Parkinson’s UK, Cure Parkinson’s Trust, PSP Association, CBD Solutions, Drake Foundation, Medical Research Council, and Michael J Fox Foundation. RR is funded by ASAP.

*KCL:* The London Neurodegenerative Diseases Brain Bank, KCL, receives funding from the MRC and as part of the Brains for Dementia Research project (jointly funded by the Alzheimer’s Society and Alzheimer’s Research UK).

*Cambridge*: Cambridge Brain Bank is supported by the NIHR Cambridge Biomedical Research Centre. JBR receives support from Wellcome Trust (220258) and NIHR Cambridge Biomedical Research Centre (BRC-1215-20014). The views expressed are those of the authors and not necessarily those of the NIHR or the Department of Health and Social Care; PSP Association and Evelyn Trust; Medical Research Council (SUAG051 R101400).

*Bristol*: We would like to thank the Southwest Dementia Brain Bank (SWDBB), their donors and donor’s families for providing brain tissue for this study. The SWDBB is part of the Brains for Dementia Research programme, jointly funded by Alzheimer’s Research UK and Alzheimer’s Society and is supported by BRACE (Bristol Research into Alzheimer’s and Care of the Elderly) and the Medical Research Council.

*Oxford*: We acknowledge the Oxford Brain Bank, supported by the Medical Research Council (MRC), Brains for Dementia Research (BDR) (Alzheimer Society and Alzheimer Research UK), Autistica UK, and the NIHR Oxford Biomedical Research Centre.

*Newcastle*: Tissue for this study was provided by the Newcastle Brain Tissue Resource which is funded in part by a grant from the UK Medical Research Council (G0400074), by NIHR Newcastle Biomedical Research Centre awarded to the Newcastle upon Tyne NHS Foundation Trust and Newcastle University, and as part of the Brains for Dementia Research Programme jointly funded by Alzheimer’s Research UK and Alzheimer’s Society.

*Manchester:* Tissue samples were supplied by The Manchester Brain Bank, which is part of the Brains for Dementia Research programme, jointly funded by Alzheimer’s Research UK and Alzheimer’s Society.

*Barcelona*: We are indebted to the HCB-IDIBAPS Biobank, integrated in the Spanish National Biobanks Network, for the biological human samples and data procurement. Gerard Piñol-Ripoll acknowledges the support from the Department of Health (PERIS 2019 SLT008/18/00050). Sergi Borrego-Écija is funded by the Joan Rodés - Josep Baselga grant from the FBBVA.

*Amsterdam*:Brain tissues were obtained from The Netherlands Brain Bank (NBB), Netherlands Institute for Neuroscience, Amsterdam (open access: www.brainbank.nl). All Material has been collected from donors for or from whom a written informed consent for a brain autopsy and the use of the material and clinical information for research purposes had been obtained by the NBB.

*Stockholm*: The Brain bank at Karolinska Institutet receives CIMED-funding.

*Paris*: The NeuroCEB Neuropathology network includes: Dr Franck Letournel (CHU Angers), Dr Marie-Laure Martin-Négrier (CHU Bordeaux), Dr Maxime Faisant (CHU Caen), Pr Catherine Godfraind (CHU Clermont-Ferrand), Pr Claude-Alain Maurage (CHU Lille), Dr Vincent Deramecourt (CHU Lille), Dr David Meyronnet (CHU Lyon), Dr André Maues de Paula (CHU Marseille), Pr Valérie Rigau (CHU Montpellier), Pr Danielle Seilhean (CHU PS, Paris), Dr Susana Boluda (CHU PS, Paris), Dr Isabelle Plu (CHU PS, Paris), Dr Dan Christian Chiforeanu (CHU Rennes), Dr Florent Marguet(CHU Rouen), Dr Béatrice Lannes (CHU Strasbourg).

*Victoria*: Brain tissues were received from the Victorian Brain Bank, supported by The Florey, The Alfred, Victorian Institute of Forensic Medicine and Coroners Court of Victoria and funded in part by Parkinson’s Victoria, MND Victoria, FightMND, Yulgilbar Foundation and Ian and Maria Cootes.

*Sydney*: JBK is supported by NHMRC Dementia Team 1095127. GMH receives funding from NHMRC program grants 1037746 and 1132524, NHMRC Dementia Team 1095127, and NHMRC Fellowships 1079679 and 1176607. OP receives funding from by NHMRC program grant 1132524 and NHMRC Dementia Team 1095127, and NHMRC Fellowships 1103258 and 2008020. JJK and JRH both receive funding from NHMRC program grants 1037746 and 1132524, and NHMRC Dementia Team 1095127.

## Author Contributions

RRV, WJS, RR, DD, JDR, JAH and OAR were involved in conceptualisation and design of study. RRV, WJS, SFR, TL, MGH and MS carried out the formal analysis including pathology review and statistical analysis. RV, WS and OAR wrote the original draft of manuscript. All authors were involved in funding acquisition, resources, validation, critically reviewing and approving final version of manuscript.

