## Supplemental Tables for "Creating the Pick’s disease International Consortium: Association study of *MAPT* H2 haplotype with risk of Pick’s disease"

| Variant | Minor allele count and frequency | Major allele count and frequency | Genotype 1 count and frequency | Genotype 2 count and frequency | Genotype 3 count and frequency |
|---|---|---|---|---|---|
| rs1467967 | | | | | |
| Pick’s disease | G: 190 (28.1%) | A: 486 (71.9%) | AA: 176 (52.1%) | AG: 134 (39.6%) | GG: 28 (8.3%) |
| Controls | G: 849 (32.4%) | A: 1775 (67.6%) | AA: 603 (46.0%) | AG: 569 (43.4%) | GG: 140 (10.7%) |
| rs242557 | | | | | |
| Pick’s disease | A: 236 (34.9%) | G: 440 (65.1%) | GG: 146 (43.2%) | GA: 148 (43.8%) | AA: 44 (13.0%) |
| Controls | A: 965 (36.8%) | G: 1659 (63.2%) | GG: 548 (41.8%) | GA: 563 (42.9%) | AA: 201 (15.3%) |
| rs3785883 | | | | | |
| Pick’s disease | A: 114 (16.9%) | G: 562 (83.1%) | GG: 231 (68.3%) | GA: 100 (29.6%) | AA: 7 (2.1%) |
| Controls | A: 472 (18.0%) | G: 2152 (82.0%) | GG: 879 (67.0%) | GA: 394 (30.0%) | AA: 39 (3.0%) |
| rs2471738 | | | | | |
| Pick’s disease | T: 136 (20.1%) | C: 540 (79.9%) | CC: 215 (63.6%) | CT: 110 (32.5%) | TT: 13 (3.8%) |
| Controls | T: 542 (20.7%) | C: 2082 (79.3%) | CC: 842 (64.2%) | CT: 398 (30.3%) | TT: 72 (5.5%) |
| rs8070723 | | | | | |
| Pick’s disease | G: 196 (29.0%) | A: 480 (71.0%) | AA: 167 (49.4%) | AG: 146 (43.2%) | GG: 25 (7.4%) |
| Controls | G: 603 (23.0%) | A: 2021 (77.0%) | AA: 784 (59.8%) | AG: 453 (34.5%) | GG: 75 (5.7%) |
| rs7521 | | | | | |
| Pick’s disease | A: 278 (41.1%) | G: 398 (58.9%) | GG: 117 (34.6%) | GA: 164 (34.6%) | AA: 57 (16.9%) |
| Controls | A: 1223 (46.6%) | G: 1401 (53.4%) | GG: 385 (29.3%) | GA: 631 (48.1%) | AA: 296 (22.6%) |

*Supplementary Table 1: Genotype counts and frequencies of six common MAPT SNPs in Pick’s disease cases and controls.*

| | Minor allele frequency | | Association with Pick's disease | |
|---|---|---|---|---|
| Variant | Pick's disease patients (N=338) | Controls (N=1,312) | OR (95% CI) | P-value |
| rs1467967 | 28.1% | 32.4% | 0.83 (0.68, 1.00) | 0.046 |
| rs242557 | 34.9% | 36.8% | 0.94 (0.79, 1.12) | 0.51 |
| rs3785883 | 16.9% | 18.0% | 0.91 (0.72, 1.15) | 0.42 |
| rs2471738 | 20.1% | 20.7% | 0.96 (0.78, 1.18) | 0.70 |
| rs8070723 | 29.0% | 23.0% | 1.35 (1.12, 1.64) | 0.0021 |
| rs7521 | 41.1% | 46.6% | 0.81 (0.69, 0.96) | 0.018 |

*Supplementary Table 2: Associations between individual MAPT variants and risk of Pick's disease. ORs, 95% CIs, and p-values result from logistic regression models that were adjusted for age and sex. ORs correspond to each additional minor allele of the given variant. OR=odds ratio; CI=confidence interval.*

| | | Association with age of disease onset | | Association with disease duration | |
|---|---|---|---|---|---|
| Haplotype | Minor allele frequency (N=309) | β (95% CI) | P-value | β (95% CI) | P-value |
| rs1467967 | 28.8% | 0.03 (-1.36, 1.41) | 0.97 | -0.11 (-0.80, 0.59) | 0.76 |
| rs242557 | 34.6% | -0.58 (-1.89, 0.72) | 0.38 | -0.42 (-1.07, 0.24) | 0.22 |
| rs3785883 | 16.8% | -0.33 (-2.05, 1.39) | 0.71 | 0.08 (-0.79, 0.94) | 0.86 |
| rs2471738 | 19.9% | -0.16 (-1.73, 1.40) | 0.84 | 0.01 (-0.77, 0.80) | 0.98 |
| rs8070723 | 29.6% | -0.54 (-1.94, 0.87) | 0.45 | 0.25 (-0.46, 0.96) | 0.50 |
| rs7521 | 40.8% | 1.11 (-0.18, 2.40) | 0.091 | -0.40 (-1.05, 0.26) | 0.23 |

*Supplementary Table 3: Associations of individual MAPT variants with age of disease onset and disease duration in Pick's disease subjects. β values, 95% CIs, and p-values result from linear regression models that were adjusted for sex and series (age of disease onset analysis) or sex, age of disease onset, and series (disease duration analysis). β values are interpreted as the change in the mean value of the given outcome (age of disease onset or disease duration) corresponding to each additional copy of the minor allele of the given variant. β=regression coefficient; CI=confidence interval.*
